# Lineage BA.2 dominated the Omicron SARS-CoV-2 epidemic wave in the Philippines

**DOI:** 10.1101/2022.05.30.22275783

**Authors:** Yao-Tsun Li, Francisco Gerardo M. Polotan, Gerald Ivan S. Sotelo, Anne Pauline A. Alpino, Ardiane Ysabelle M. Dolor, Ma. Angelica A. Tujan, Ma. Ricci R. Gomez, Othoniel Jan T. Onza, Angela Kae T. Chang, Criselda T. Bautista, June C. Carandang, Dodge R. Lim, Lei Lanna M. Dancel, Mayan Uy-Lumandas, Timothy John R. Dizon, Katie Hampson, Simon Daldry, Joseph Hughes, Kirstyn Brunker

## Abstract

The Omicron SARS-CoV-2 variant led to a dramatic global epidemic wave following detection in South Africa in November, 2021. The Omicron lineage BA.1 was dominant and responsible for most domestic outbreaks during December 2021-January 2022, whilst other Omicron lineages including BA.2 accounted for the minority of global isolates. Here, we describe the Omicron wave in the Philippines by analysing genomic data. Our results identify the presence of both BA.1 and BA.2 lineages in the Philippines in December 2021, before cases surged in January 2022. We infer that only lineage BA.2 underwent sustained transmission in the country, with an estimated emergence around November 18th, 2021 [95% highest posterior density: November 6-28th], whilst despite multiple introductions BA.1 transmission remained limited. These results suggest the Philippines was one of the earliest areas affected by BA.2, and reiterate the importance of whole-genome sequencing for monitoring outbreaks.

## Introduction

The continuous transmission of severe acute respiratory syndrome coronavirus 2 (SARS-CoV-2), the aetiology of the coronavirus disease (COVID-19), has led to new viral variants with accumulated genetic mutations (Rambaut et al. 2020; Harvey et al. 2021). The Omicron variant was designated by the WHO as a Variant of Concern (VOC) in November, 2021, following previously designated Alpha, Beta, Gamma and Delta VOCs (WHO 2021a). These SARS-CoV-2 variants each possess distinct combinations of mutations in the viral genome, particularly in the S gene, demonstrating potential for increased transmissibility or disease severity compared with viruses isolated early in the pandemic (Dhar et al. 2021; Volz et al. 2021; Viana et al. 2022). VOCs may circulate efficiently in the population by evading antibodies derived from vaccination or prior exposures, or if they elude diagnostic methods (Dhar et al. 2021; Volz et al. 2021; Liu et al. 2022; Schmidt et al. 2022). Thus effectively tracking the emergence and evolution of SARS-COV-2 lineages is essential to controlling the disease.

The Omicron variant was first reported in South Africa in October, 2021 (Viana et al. 2022), with three divergent lineages identified (BA.1, BA.2 and BA.3). Of the three lineages, the lineage BA.1 (including its descending sublineages BA.1.*) has rapidly spread to dominate globally, leading to another epidemic wave during the 2021 winter (Hodcroft 2021; WHO 2021b; Chen et al. 2022). In contrast, the BA.2 and BA.3 lineages had only accounted for a minority of viral isolates by the end of 2021 (Hodcroft 2021; Chen et al. 2022). The BA.2 viruses, although phylogenetically clustered with BA.1 compared to other variants (Viana et al. 2022; Yamasoba et al. 2022), differ by at least 30 amino acids relative to BA.1 viruses (Majumdar and Sarkar 2022; Tsueng et al. 2022). Recent studies provide hints that significant genetic divergence of the two lineages result in different replication capacity and transmissibility (Lyngse et al. 2022; UKHSA 2022; Yamasoba et al. 2022).

The Philippines was among a few countries, including Denmark and India, where the BA.2 lineage accounted for noticeable genomic data during the 2021 winter, contrary to most geographical areas mainly affected by the BA.1 lineage (Colson et al. 2022; Desingu and Nagarajan 2022). The country also experienced a sharp increase in case numbers in January, 2022, parallel to the global Omicron wave (Department of Health, Philippines); nevertheless, lineages contributing to local transmission and their dynamic interactions were unknown. In this study, we show using phylogenetic approaches that the BA.2 lineage but not the BA.1 lineage caused the case surge in the Philippines during the global Omicron wave. We also inferred that BA.2 circulation in the Philippines could have occurred as early as November, 2021, within weeks of the lineage first being identified in South Africa. These results provide insights into how new SARS-CoV-2 lineages emerge and establish sustained transmission.

## Materials and Methods

### Genomic surveillance in the Philippines

SARS-CoV-2 sequences were collected under the framework of a collaborative project, GECO (Genomic Epidemiology of COVID in the Philippines). The project aims to use viral genomes generated by the nanopore sequencing to inform public health measures against COVID-19. SARS-CoV-2 PCR-positive RNA samples from partnered Sub-National Laboratories (SNLs) were subjected to whole genome sequencing using the ARTIC network multiplex PCR workflow performed at a national core laboratory, the Research Institute for Tropical Medicine (RITM). The ARTIC network bioinformatic pipeline was used to generate consensus sequences from raw output files with steps of basecalling, de-multiplexing, mapping and polishing (https://artic.network/ncov-2019). As of February 15th, 2022, 1055 consensus sequences of SARS-CoV-2 have been generated by the project.

### Sequence data preparation

To compile all available genomic data from the Philippines and fit the domestic isolates in the context of global virus transmission, all SARS-CoV-2 sequences and metadata were downloaded from GISAID on February 15th, 2022 (EpiCoV, https://www.gisaid.org). The downloaded data were first split into Philippine/non-Philippine portions based on the location of isolation, in which the Philippine data deposited in GISAID were then combined with data collected by the GECO project. An Omicron data set containing all Omicron sub-lineages from the Philippines and a BA.2 lineage data set containing only BA.2 lineage data from the country were prepared according to the Pango lineages (Rambaut et al. 2020) assigned to each sequence by Pangolin version 2022-02-02 (https://github.com/cov-lineages/pangolin) or information provided by GISAID. For each data set, 1500 global proximal strains genetically similar to the Philippine strains were sampled by the Nextstrain bioinformatic pipeline (Hadfield et al. 2018). The quality of the compiled genomic data was evaluated by Nextclade CLI v1.10.3 (Aksamentov et al. 2021). We filtered out sequences that had more than 5 private mutations or a SNP cluster. Sequences shorter than 27000 nucleotides or sequences excluded by Nextclade due to too many ambiguous sites were also removed from the data sets. Accession numbers of the sequences analysed in this study have been compiled as an EPI SET (EPI_SET_20220430vo).

### Phylogenetic and other genetic analyses

Curated whole genome SARS-CoV-2 sequences were aligned using Nextalign v1.9 (Aksamentov et al. 2021), and the alignments supplemented with a reference strain Wuhan/Hu-1/2019 were subject to maximum likelihood (ML) tree inference using IQ-TREE v2.2.0 (Minh et al. 2020). To focus on domestic transmission in the Philippines and cross-border events, we subsampled the compiled Omicron data set by selecting a taxon in each monophyletic group that comprised only strains isolated from the same country outside the Philippines, and the reduced data set was then used to rebuild another ML tree. Based on the resulting ML tree, the time-scaled tree of the Omicron variant was estimated using TreeTime v0.8.5 with a clock rate of 0.0008. Times of the most recent common ancestor (tMRCAs) of specific taxa can be parsed from the internal nodes in the time-scaled tree.

To more closely explore the timing of the BA.2 introduction, a Bayesian phylogenetic framework was implemented with BA.2 sequences collected in the Philippines. A more strictly filtered BA.2 data set was prepared with sequences annotated as good quality by Nextclade. With this, BA.2 genome data for the time-calibrated phylogeny subsequently included all filtered Philippine BA.2 sequences isolated before January 15th, 2022, with apparently divergent BA.2 strains removed (n=19), and early BA.2 strains in South Africa and India. A time-scaled phylogeny was inferred using BEAST v10.4 (Suchard et al. 2018) facilitated by the BEAGLE library v3.1 for better computational performance (Ayres et al. 2019). We employed a HKY plus gamma substitution model, and a strict molecular clock with an exponential demographic prior in the Bayesian analyses. Markov Chain Monte Carlo (MCMC) analysis was run for 100 million steps and sampled every 10,000 steps. Three parallel runs were performed and combined with a burnin of 10 million per chain using LogCombiner (Suchard et al. 2018). Parameters logged during the MCMC runs were inspected by Tracer v1.7.1 (Rambaut et al. 2018). A summarized maximum clade credibility (MCC) tree was inferred using TreeAnnotator (Suchard et al. 2018).

Genetic divergence was calculated by sequence length times genetic diversity (pi)(Nei et al. 2000). Introductory events and the local clusters were identified using clusterfunk v0.1.0 (https://github.com/snake-flu/clusterfunk) with phylogenetic trees inferred by IQ-TREE. Statistical correlation between locations of isolation and the phylogeny was detected by BaTS v0.9 with 1000 posterior trees subsampled from the MCMC process (Parker et al. 2008). All phylogenetic trees were visualized by ggtree (Yu et al. 2017).

## Results

The epidemic wave associated with the Omicron variant in the Philippines started in December, 2021. Based on case information available from the Department of Health, Philippines, reported COVID-19 case numbers rose towards the end of 2021, reaching a peak with over 30,000 cases per day in week 2 (10 January-16 January), 2022, before rapidly declining to fewer than 5,000 cases per day in week 6 (7 February-13 February)(Figure 1A, barchart). Numbers of cases identified from returning overseas Filipinos (ROFs) demonstrate a remarkably similar epidemic profile to the reported domestic cases (Figure 1A, line), suggesting linkage of global SARS-CoV-2 transmission to the domestic epidemic.

**Figure 1.**
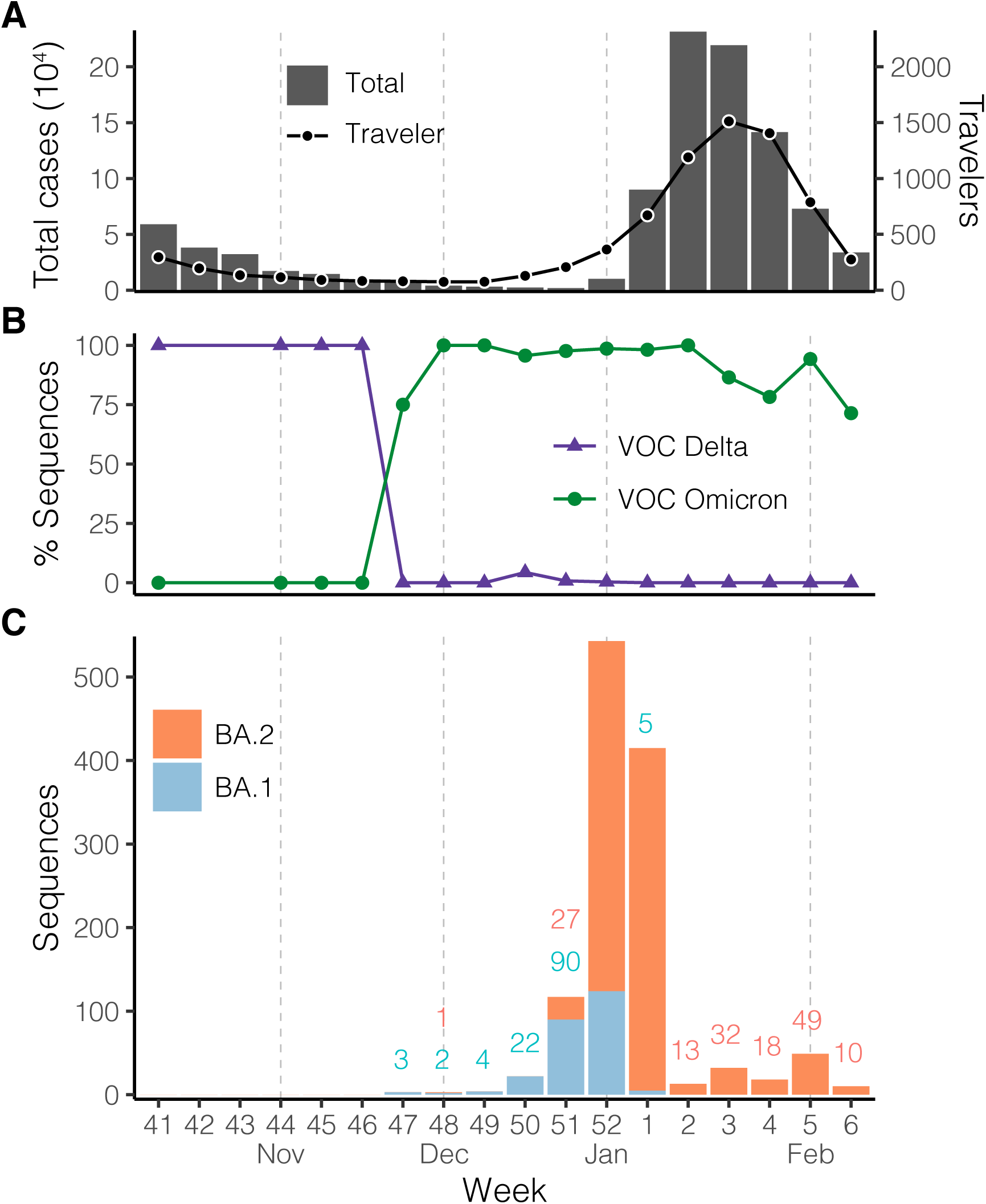
Severe acute respiratory syndrome coronavirus 2 (SARS-CoV-2) infections during the Omicron variant epidemic wave in the Philippines. (A) Total numbers of cases reported (barchart) in the country and numbers of cases identified from returning overseas Filipinos (line) based on the Case information Data from the Department of Health, Philippines. (B) Proportions of variant sequences among available SARS-CoV-2 sequences as of February 15, 2022. The classification was based on the software Pangolin version 2022-02-02. (C) Numbers of available Omicron lineage sequences as of February 15, 2022. Numbers below 100 are annotated (coloured by lineage) above the bars. BA.1 and BA.2 categories here contain the descending lineages assigned by Pangolin, e.g. BA.1 includes the lineage BA.1.1. X-axis labels indicate epidemiological week defined by the US-CDC, which corresponds to the ISO week starting on a Sunday. VOC, Variant of Concern.

To better understand the transmission of SARS-CoV-2 viruses leading to the case surge, we combined sequence data collected by the GECO project (Genomic Epidemiology of COVID in the Philippines) and data available on the GISAID. The sequences in the Philippines show the Delta variant, including lineages 1.617.2 and AY.*, as being the dominant circulating variant in the country before being replaced by the Omicron variant (lineages BA.*)(Figure 1B). Specifically, the proportion of sequenced cases belonging to the Omicron variant exceeded the Delta variant in November, 2021, about one month before the rise of the epidemic wave (Figure 1A and 1B), and the Omicron variant has accounted for the majority of sequences since. Among the Omicron variant viruses in the Philippines, BA.1 lineage, first identified on 22 November, had accounted for more available sequences than its sister lineage BA.2 until the last week of 2021 (Figure 1C), although the numbers of both lineages were low at most time points during November-December, 2021. In contrast, BA.2 has been the most prevalent since the lineage drastically increased in the last week of the year 2021 (Figure 1C).

BA.1 and BA.2 viruses isolated in the Philippines show divergent distributions on the phylogeny inferred by the whole viral genome. With global strains sampled in an unbiased manner against the proximal Philippines isolates, the BA.1 viruses isolated in the Philippines are intermixed with the non-Philippine viruses on the temporal phylogenetic tree suggesting a large number of introductions. In contrast, BA.2 viruses are largely clustered in one clade, in which the most genetically similar virus of each Philippine isolate is nearly always from the Philippines (Figure 2A). Estimation of introductory events by ancestral state reconstruction using the parsimony method also supports this observation: 136 potential introductory events to the Philippines were identified in the BA.1 lineage, which led to clusters with a mean sample size below 2, compared with 25 potential introductory events identified in the BA.2 lineage, which led to two major clusters with sizes of 699 and 206 in addition to the remaining clusters each having less than 10 samples.

**Figure 2.**
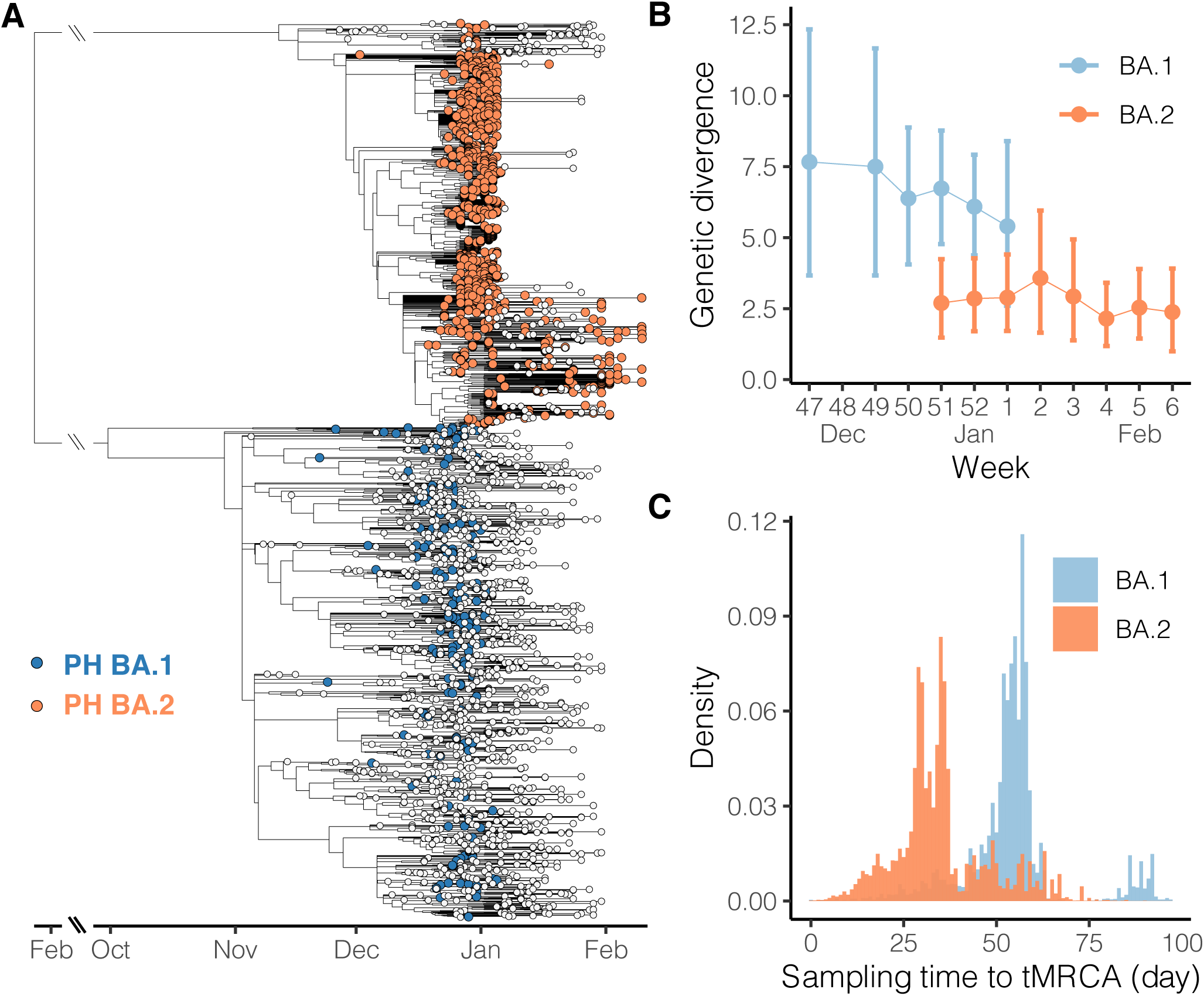
Phylogenetic relationship of SARS-CoV-2 Omicron variants isolated in the Philippines. (A) Time-scaled tree was inferred by TreeTime using Omicron variant genome sequences isolated in the Philippines and from the global database. Blue tips and orange tips indicate BA.1 and BA.2 lineage viruses, respectively, isolated in the Philippines, whereas white tips indicate viruses isolated in the other countries. Long branches descending from the common ancestor of BA.1 and BA.2 are shortened. (B) Average genetic divergence of viruses isolated in the Philippines. Error bars represent 95% bootstrap percentiles. (C) Distribution of time intervals from time of the most common ancestor (tMRCA) to the isolation time. The time intervals were calculated based on pairs of Philippine taxa on the time-scaled tree (panel A); for each pair the larger time difference was recorded.

We therefore hypothesise that the two Omicron lineages in the Philippines demonstrate different epidemiological patterns. We assume if most genomic samples were collected from the context of sustained transmission rather than sporadic introduction, genetic differences between sequences isolated in approximate time points would be minimal. Additionally, if most samples were collected from sustained transmission, viral taxa would coalesce to close common ancestors on the phylogeny, in contrast to deeper common ancestors likely shared by taxa isolated in unlinked transmission chains. Our results show BA.1 lineage sequences grouped by week have greater average genetic differences compared with BA.2 lineage sequences (Figure 2B). Especially, among the 2 or 3 weeks where both lineages are available, average nucleotide differences shared by paired BA.1 sequences are more than twice that of the BA.2 sequences. Although the BA.2 lineage in the Philippines has more overall samples than the BA.1 lineage (Figure 1C), weekly genetic differences of BA.2 remain stable throughout the studied intervals, averaging about 2.5 nucleotide differences among sequences isolated in each week (Figure 2B). Furthermore, when pairwise time intervals between the most recent common ancestor and isolation time are compared between the two lineages, BA.1 lineage shows greater intervals than BA.2 lineage (Figure 2C). The average time intervals of lineage BA.1 and BA.2 are 54 and 35 days, respectively, indicating each pair of BA.1 viruses have deeper common ancestors than the BA.2 viruses. Combined with the observations where no distinguishable Philippines clade was formed among the BA.1 global isolates (Figure 2A), these comparative analyses suggest that the BA.2 but not BA.1 lineage underwent sustained transmission in the Philippines.

To gain more insights to the introduction of the BA.2 lineage in the Philippines, we estimated the time of most recent common ancestor (tMRCA) based on the BA.2 genomic sequences isolated in the country in addition to early BA.2 strains identified globally (Figure 3). The estimated tMRCA is the18 November, 2021 (95% highest posterior density (HPD), 6 November-28 November), two weeks before the first BA.2 case identified in the Philippines. The estimated tMRCA does not significantly differ from the root of the BA.2 temporal phylogeny (95 HPD, 23 October-17 November), which may corroborate with the previous understanding that the Philippines was one of the earliest countries where circulation of the BA.2 lineage was discovered. No apparent diffusion pattern was observed based on the geographical distribution on the phylogeny (Figure 3). Viruses isolated from the three major island groups are generally mixed on the subclades, providing evidence of extensive domestic transmission and underlying circulation before increased sampling in January. Indeed, available sequences assigned as BA.1 have only been isolated from 9 administrative regions, compared with BA.2 isolated from 16 regions (Supplementary Figure). Among these regions, early BA.2 from the National Capital Region (Luzon island group), Ilocos (Luzon) and the Eastern Visayas (Visayas) show statistically significant clustering on the phylogeny (Supplementary Table).

**Figure 3.**
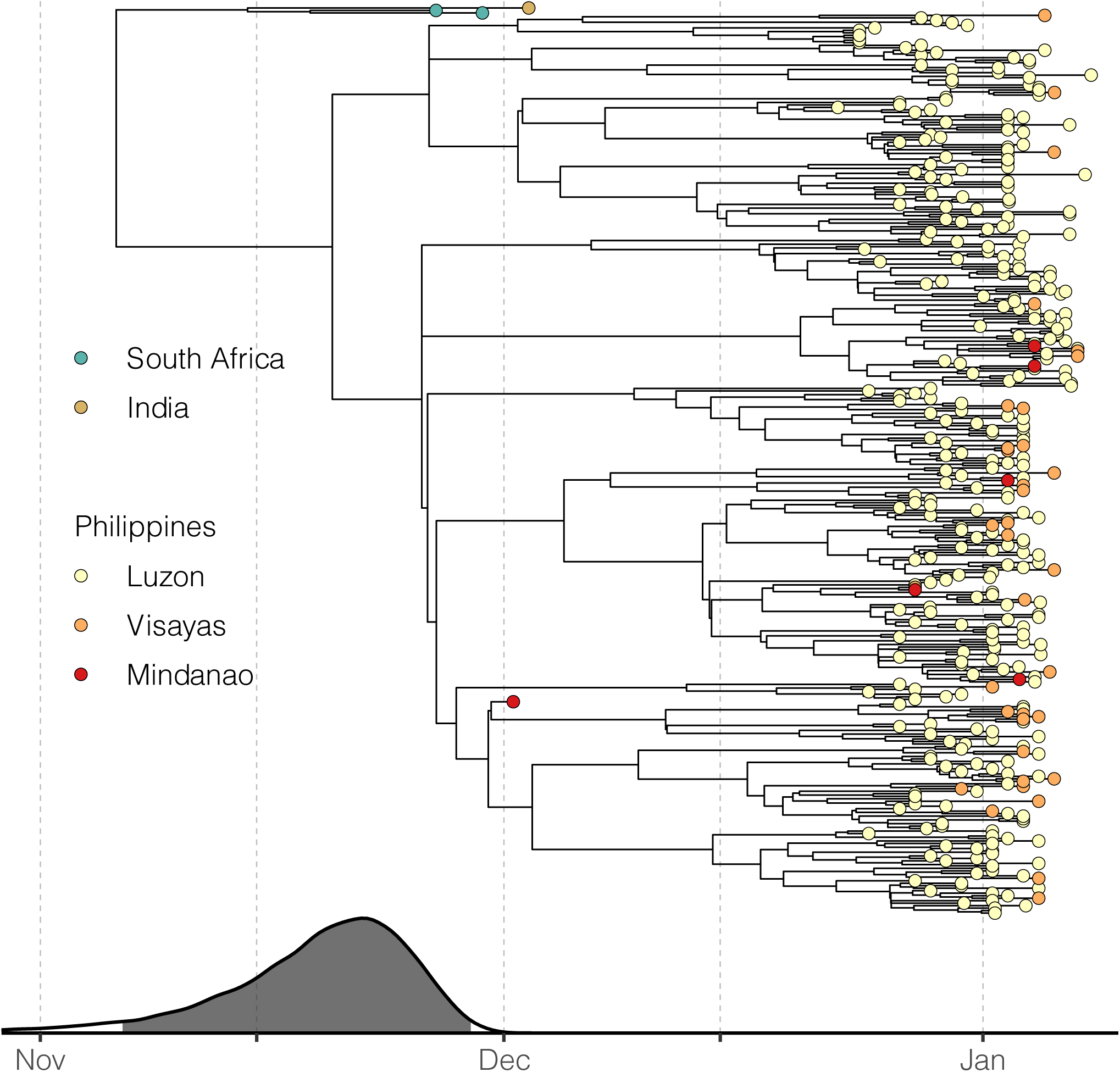
Introduction of BA.2 lineage in the Philippines. Time-scaled tree was inferred by BEAST using BA.2 genomes isolated in the Philippines along with genomes of early global BA.2 viruses. Estimated tMRCAs with 95% highest posterior density (HPD) illustrated by gray area are aligned with the phylogenetic tree. Tips are colored according to the location of isolation. Red shades for viruses isolated in the Philippines indicate the three island groups in the country.

## Discussion

In this study, we show that the BA.2 but not the BA.1 lineage of the Omicron variant fit the scenario of community transmission in the Philippines based on viral genomic data. With the majority of sequences isolated during the country’s latest case rise identified as the BA.2 lineage, we propose the Omicron epidemic wave in the Philippines was mostly driven by BA.2 viruses in contrast to most other Omicron waves seen during this time globally. In most countries with continuous genomic surveillance, including South Africa, UK and the USA, case peaks associated with the Omicron variant during the 2021 winter were caused by the BA.1 (including descendant BA.1.*) lineage (Chen et al. 2022; Tsueng et al. 2022). In contrast, the BA.2 lineage was only observed to be dominant in a few countries besides the Philippines, including India, Nepal, Bangladesh, Denmark and Qatar, by the end of January 2022 based on the available genomic data (Hodcroft 2021; Chen et al. 2022). Our results in the Philippines thus present a special case whereby the BA.2 lineage led to local transmission without a previous extensive BA.1 outbreak.

Since March 2022, there have been clear signs that BA.2 has replaced BA.1 in several geographical regions (Chen et al. 2022; Tsueng et al. 2022). Understanding how the BA.2 lineage became dominant over the previous circulating variants in the Philippines could provide important insights for controlling BA.2 in affected countries. As there appears to have been only a low level of local BA.1 circulation in the Philippines, it is not directly clear whether virological properties of the two Omicron lineages, including intrinsic transmissibility or antigenicity (Lyngse et al. 2022; Schmidt et al. 2022; UKHSA 2022; Yamasoba et al. 2022), competitively determined the epidemic outcome through selection of a more fit strain. Since Omicron emerged, routine testing using the S-gene target failure (SGTF) marker has been implemented to detect and curtail the spread of BA.1 by distinguishing it from the Delta variant. However, this method identifies BA.1 but rarely BA.2 based on the deletions in the viral S gene (Majumdar and Sarkar 2022; UKHSA 2022), and therefore may have led to reduced detection of BA.2, potentially favouring the emergence and spread of BA.2 viruses. This point emphasises the importance of whole genome sequencing as part of SARS-CoV-2 surveillance programmes.

We estimated the most recent common ancestor of the Philippine BA.2 viruses to be in late November, 2021, about 2 weeks before the first detected case of BA.2 in the Philippines. This estimate by Bayesian phylogenetic reconstruction is corroborated with the time-scaled tree inferred using the maximum likelihood method (Figure 2A, the tMRCA of the major clade formed by Philippine taxa), suggesting local spread could have started in November. The BA.2 lineage was first described in South Africa in early November, 2021, along with other Omicron lineages (Viana et al. 2022). Local BA.2 transmission was then reported in Denmark where the first BA.2 case was identified on 5 December, 2021 (Fonager et al. 2022), and the country accounted for most global BA.2 sequences as of mid-January 2022 (>5000)(Desingu and Nagarajan 2022). India was also identified as one of the earliest BA.2 affected countries (Colson et al. 2022; Desingu and Nagarajan 2022), with BA.2 sequences isolated as early as November, 2021 (Hodcroft 2021). The temporal phylogeny estimated from our BA.2 dataset ascertained multiple introductory events, based on the topology of the tree and limited sampling in early December (Figure 3). Despite uncertainty in the exact origin of currently circulating BA.2 viruses in the Philippines, the estimated date of emergence appears robust to sampling effects. The two BA.2 subclades, either with or without the first isolate on the 3rd of December, could still trace the most recent common ancestor’s origin before December (Figure 2A and 3).

The estimated BA.2 emergence time coincides with the general de-escalation of control measures in the Philippines. The de-escalation may be attributed to the decreasing number of identified COVID-19 infections in November, 2021. At this time new COVID-19 infections reached their lowest number for the previous 11 months (Department of Health, Philippines). There was also general optimism and anticipation for the Christmas season, during which migrant Filipino workers come home to celebrate with their families. How migrant workers contributed to local transmission warrants further research.

Variation in sequencing rates across administrative regions in the Philippines render our genomic data unlikely to reflect the domestic geographical diffusion of lineages in the Philippines. Our comparative analyses between BA.1 and BA.2 lineages, nevertheless, are less affected by undersampling since the analyses were based on the topology of the phylogeny and the sample selection for sequencing would not be biased by lineage. Importantly, routine genomic surveillance in the Philippines shows no evidence of emergence of the BA.1 lineage (https://geco-ph.github.io/GECO-covid). Retrospective sequencing of Philippine and global samples will facilitate improved understanding of the BA.2 origin and reconstruction of viral diffusion dynamics during the pandemic. The root of the temporal phylogeny of BA.2 is estimated at 5 November, which is very close to the emergence date estimated by a recent study (6 November)(Yamasoba et al. 2022).

In summary, we show the epidemic wave in the Philippines was driven by Omicron lineage BA.2 but not BA.1, although both lineages were sampled before and during the rise in case numbers. Also, the BA.2 viruses causing the country’s epidemic circulated in the Philippines before December, 2021, in parallel to Denmark as one of the earliest countries where local BA.2 outbreaks occurred. Our study highlights the value of phylogenetic methods for understanding viral transmission, and the need to rapidly generate genomic data to inform control strategies.

## Data Availability

Genomic data collected by the GECO project are available on GISAID, and accession numbers of the sequences analysed in this study have been compiled as an EPI SET (EPI_SET_20220430vo). The XML file required for BEAST, details of genetic analyses including phylogenetic trees are available at https://github.com/GECO-PH/GECO-covid/tree/main/manuscript_BA2.

## Acknowledgements

The work is supported by the UK Research and Innovation Global effort on COVID-19 (MR/V035444/1) and Wellcome (207569/Z/17/Z).

## Supplementary data

**Supplementary Table.** Correlation between geographical location and phylogeny.

| Region | Observed mean<br>(95% CI) | Null mean<br>(95% CI) | Significance |
| --- | --- | --- | --- |
| **Global** | 2.36<br>(1-3) | 1.00<br>(1-1.02) | 0.005 |
| **National Capital**<br>**Region** | 7.47<br>(6-11) | 6.15<br>(5.68-6.84) | 0.04 |
| Central Luzon | 2.07<br>(2-3) | 1.48<br>(1.28-2.01) | 0.06 |
| Cagayan Valley | 1.03<br>(1-1) | 1.04<br>(1-1.09) | 1.0 |
| Calabarzon | 2.60<br>(2-4) | 2.23<br>(2.06-2.44) | 1.0 |
| Western Visayas | 1.15<br>(1-2) | 1.17<br>(1.07-1.40) | 1.0 |
| Cordillera Administrative<br>Region | 1.01<br>(1-1) | 1.01<br>(1.0-1.03) | 1.0 |
| **Ilocos** | 2.18<br>(2-3) | 1.11<br>(1.03-1.21) | 0.015 |
| Soccsksargen | 1.0<br>(1-1) | 1.00<br>(1.0-1.01) | 1.0 |
| **Eastern Visayas** | 3.0<br>(3-3) | 1.14<br>(1.05-1.33) | 0.005 |
| Davao Region | 1.02<br>(1-1) | 1.01<br>(1.0-1.03) | 1.0 |
| Central Visayas | 1.07<br>(1-2) | 1.03<br>(1.00-1.08) | 1.0 |
| Mimaropa | 1.0<br>(1-1) | 1.0<br>(1.0-1.0) | 1.0 |

**Supplemnentary Figure.**
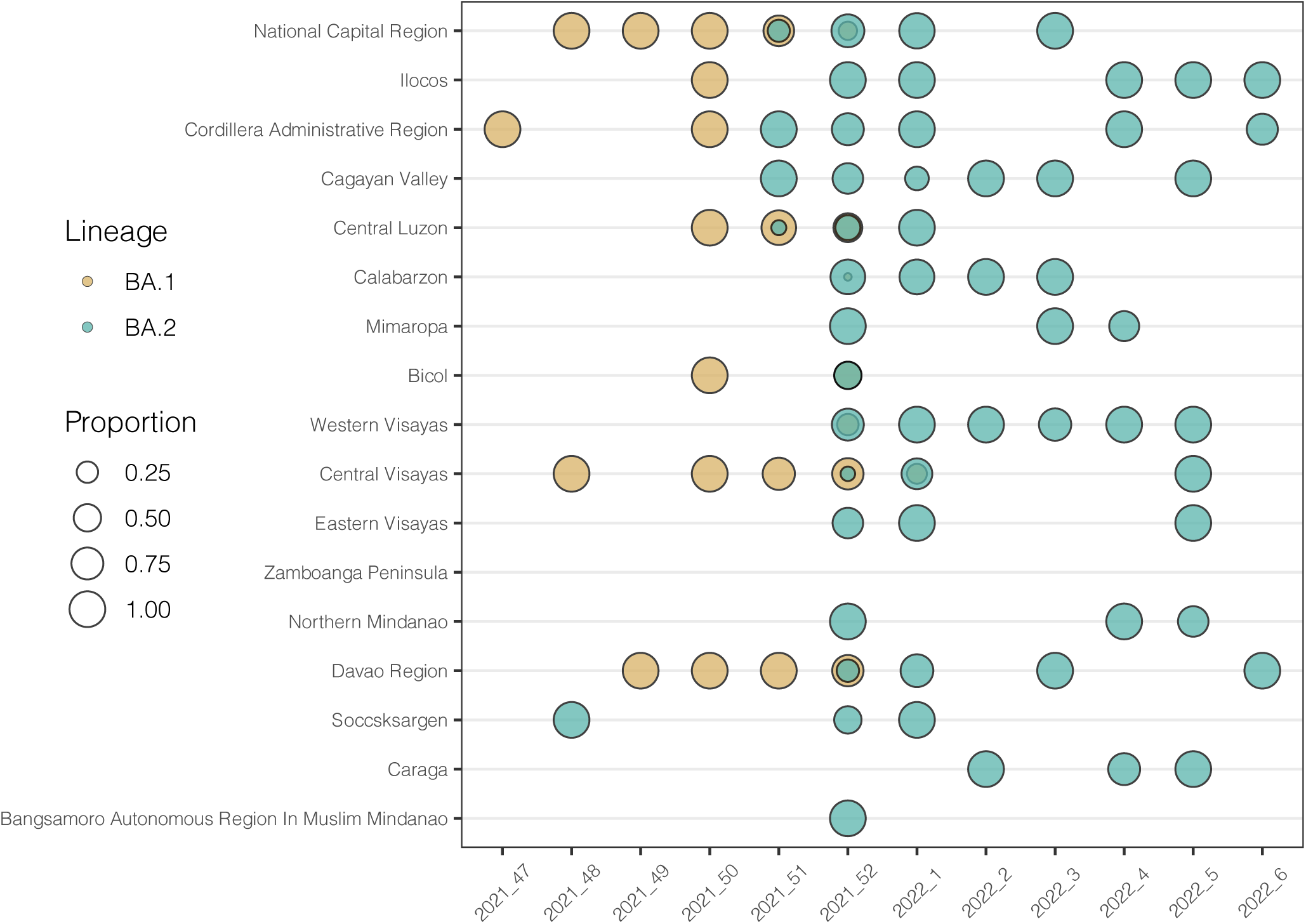
Visualisation of locations of BA.1/BA.2 samples against time. X-axis labels represent year plus week and circle sizes are scaled to the proportion of the lineage per location per week. Data source is identical to Figure.1C.

